# CWAS-Plus: Estimating category-wide association of rare noncoding variation from whole-genome sequencing data with cell-type-specific functional data

**DOI:** 10.1101/2024.04.15.24305828

**Authors:** Yujin Kim, Minwoo Jeong, In Gyeong Koh, Chanhee Kim, Hyeji Lee, Jae Hyun Kim, Ronald Yurko, Il Bin Kim, Jeongbin Park, Donna M. Werling, Stephan J. Sanders, Joon-Yong An

**Affiliations:** Department of Integrated Biomedical and Life Science, Korea University, Seoul, 02841, Republic of Korea; L-HOPE Program for Community-Based Total Learning Health Systems, Korea University, Seoul, 02841, Republic of Korea; School of Biosystem and Biomedical Science, College of Health Science, Korea University, Seoul, 02841, Republic of Korea; Department of Statistics and Data Science, Carnegie Mellon University, Pittsburgh, PA, 15213, USA; Department of Psychiatry, CHA Gangnam Medical Center, CHA University School of Medicine, Seoul, 06135, Republic of Korea; School of Biomedical Convergence Engineering, Pusan National University, Busan, 50612, Republic of Korea; Laboratory of Genetics, University of Wisconsin-Madison, Madison, WI, 53706, USA; Institute of Developmental and Regenerative Medicine, Department of Paediatrics, University of Oxford, Oxford, OX3 7TY, UK; Department of Psychiatry and Behavioral Sciences, UCSF Weill Institute for Neurosciences, University of California, San Francisco, CA 94158, USA

**Keywords:** genetic association, whole-genome sequencing, regulatory noncoding variant, rare variant, noncoding genome

## Abstract

Variants in cis-regulatory elements link the noncoding genome to human brain pathology; however, detailed analytic tools for understanding the association between cell-level brain pathology and noncoding variants are lacking. CWAS-Plus, adapted from a Python package for category-wide association testing (CWAS) employs both whole-genome sequencing and user-provided functional data to enhance noncoding variant analysis, with a faster and more efficient execution of the CWAS workflow. Here, we used single-nuclei assay for transposase-accessible chromatin with sequencing to facilitate CWAS-guided noncoding variant analysis at cell-type specific enhancers and promoters. Examining autism spectrum disorder whole-genome sequencing data (n = 7,280), CWAS-Plus identified noncoding *de novo* variant associations in transcription factor binding sites within conserved loci. Independently, in Alzheimer’s disease whole-genome sequencing data (n = 1,087), CWAS-Plus detected rare noncoding variant associations in microglia-specific regulatory elements. These findings highlight CWAS-Plus’s utility in genomic disorders and scalability for processing large-scale whole-genome sequencing data and in multiple-testing corrections. CWAS-Plus and its user manual are available at https://github.com/joonan-lab/cwas/ and https://cwas-plus.readthedocs.io/en/latest/, respectively.

**KEY POINTS:**

- CWAS-Plus efficiently identifies noncoding associations in WGS data, supporting user-friendly categorization and burden enrichment tests.
- CWAS-Plus integrates various functional datasets, emphasizing cell-type-specific noncoding associations.
- CWAS-Plus provides a novel approach for multiple testing correction, enhancing the reliability of the results.
- Autism spectrum disorder risk noncoding variants are identified as enriched with transcription factors, suggesting their role in the pathology.
- Rare variant analysis with Alzheimer’s disease samples reveals strong association with microglia, supporting the reliability of the results produced by CWAS-Plus.

## INTRODUCTION

The noncoding regions of the genome harbor numerous key regulatory elements involved in cellular and organ development and function. Among these, promoters and enhancers play a critical role during early human development, as they contribute to tissue-specific gene expression^1,2^. Recent large-scale single-cell assay for transposase-accessible chromatin with sequencing (ATAC-Seq) studies have identified cell type-specific noncoding elements at critical developmental milestones and profiled key regulatory enhancers of gene expression dynamics^3,4^. Disruption of these elements by genetic variants may alter gene expression in biological pathways and consequently lead to human disorders^5–7^. Notably, most of the associations from genome-wide association studies (GWAS) are in noncoding regions enriched for regulatory elements^8,9^, highlighting the importance of identifying variants within these key regulatory elements in the noncoding sequences to gain critical insights into genetic disorders.

Recent advances in whole-genome sequencing (WGS) have facilitated the identification of rare variants in the noncoding genome and the evaluation of risk stemming from noncoding variants for disorders^10,11^. Several methods have been developed to prioritize high-risk noncoding variants using a scoring system based on multiple functional annotations^12–19^. These methods assess the pathogenicity of individual rare variants in the noncoding genome but do not provide statistical evaluation for genome-wide association. With the growing volume of WGS data, genome-wide evaluation methods have been proposed for genetic association study for noncoding variants^20,21^. These methods seek to identify noncoding association for functional regions^20^ or pre-defined genomic windows^21^. Phenotype-genotype association studies with combinations of functional regions were also introduced^22^. Recently, we introduced the category-wide association study (CWAS), a novel statistical framework to identify noncoding association from WGS data^23^. This method utilizes variant groups, called categories, for genetic association testing, and creates multiple categories based on genomic and functional annotations related to noncoding variants. With this, we have successfully identified that noncoding variants in conserved regions of promoters are significantly enriched for autism spectrum disorder (ASD)^24^.

Herein, we introduce CWAS-Plus, a Python package that enhances the functionality of CWAS and further develops the CWAS framework to improve the identification of genetic association of noncoding variants. CWAS-Plus significantly reduces run time and computational requirements, resulting in faster and more efficient execution of the entire CWAS workflow. CWAS-Plus integrates multiple datasets to define functional regions or genes of interest provided by the users as bed files or gene sets. Additionally, it introduces a novel approach for multiple-testing corrections, further enhancing the reliability of the results. We applied CWAS-Plus to large-scale WGS datasets and, through the integration of single-cell datasets, found that ASD noncoding association disrupts transcriptional regulation. Furthermore, applying CWAS-Plus to rare variants in case-control WGS data identified of microglia-specific noncoding association with Alzheimer’s disease (AD). Taken together, we highlight the potential of CWAS-Plus for discovering noncoding risks from WGS data across various disorders.

## METHODS

### Input requirement

CWAS-Plus requires a list of variants and samples for association testing. For the variant list, Variant Call Format (VCF) is used to include genomic position, reference allele, alternate allele, and sample identification (ID). A sample list should contain sample IDs and phenotypic labels (e.g., case or control). Users can provide adjustment factors to correct for confounding factors. These inputs are subject to variant annotation for genomic regions by the Variant Effect Predictor (VEP)^25^.

### Annotation

The variant annotation process involves two steps: the VEP annotation and customized annotation. VEP annotates the most severe consequence using Sequence Ontology (SO) terms, which is then employed for categorizing genomic regions. In this study, VEP version 110 was used. Customized annotation is carried out for user-provided datasets, including functional annotation and functional score. Functional annotation refers to specific genomic regions associated with particular functions, such as epigenetic statuses. Functional scores indicate regions with score metrics related to genomic features such as conservation.

### Categorization

To assess the noncoding association, CWAS-Plus creates categories for variants using genomic and functional annotations. A single category is built by a combination of five features: 1) variant type, 2) genomic region, 3) gene set, 4) functional annotation, and 5) functional score (see ‘Annotation datasets used in CWAS analysis’ section in **Supplementary Materials** for datasets used in each feature).

1. Variant type: variants are classified to either single nucleotide variant (SNV) or insertion-deletion (indel) based on the length of the alleles.
2. Genomic region: genomic region based on the location of the variant relative to genes. Genomic region is defined by utilizing SO terms from the most severe consequence and the annotated gene from VEP annotation. Genomic region includes coding domain (e.g., protein-truncating variants (PTVs), frameshift indels, missense, damaging missense, in frame indels, silent variants) and noncoding domain (e.g., such as composed of 5′ UTR, 3′ UTR, promoter, splice site, intron, intergenic, long noncoding RNA, and others), with the order corresponding to the order of variant annotation. PTVs (nonsense and frameshift) must have a high confidence level (“HC”) by the LOFTEE plugin and be annotated with either ‘SINGLE_EXON’ or no LOFTEE flags. Damaging missense variants are required to have an MPC score ≥2. Promoter variants are annotated as 2,000 base pairs upstream of transcription start sites.
3. Gene set: disease-relevant gene sets. Datasets should be in text format.
4. Functional annotation: functional regions associated with epigenetic modifications or regulatory elements. Datasets should be in bed format.
5. Functional score: score metrics related to specific genomic features, such as conservation or pathogenicity. Datasets should be in bed format.

By combining five annotations, a category, such as intergenic SNVs near CHD8 target genes on excitatory neuron-specific cis-regulatory elements (CRE) and conserved loci, is created.

### Burden test

CWAS-Plus assesses the association within a single category by conducting burden tests through two approaches: variant-level and sample-level tests.

In variant-level test, the case-control association is estimated by comparing the number of variants in each category. The relative risk (RR) is calculated by the following equation:

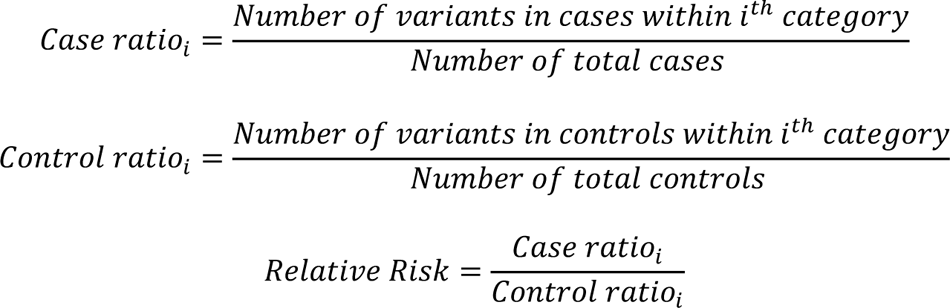

The binomial p-value is calculated by comparing two proportions: variants in cases from total variants within a category and cases from total samples.

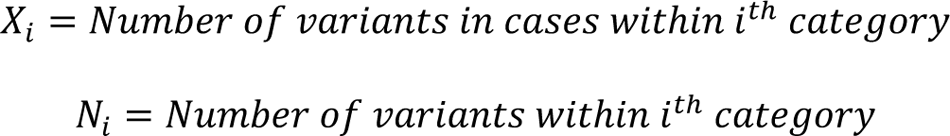

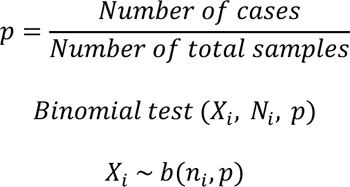

In sample-level test, the case-control association is estimated by comparing the number of samples carrying variants in each category. The RR is calculated by the following equation:

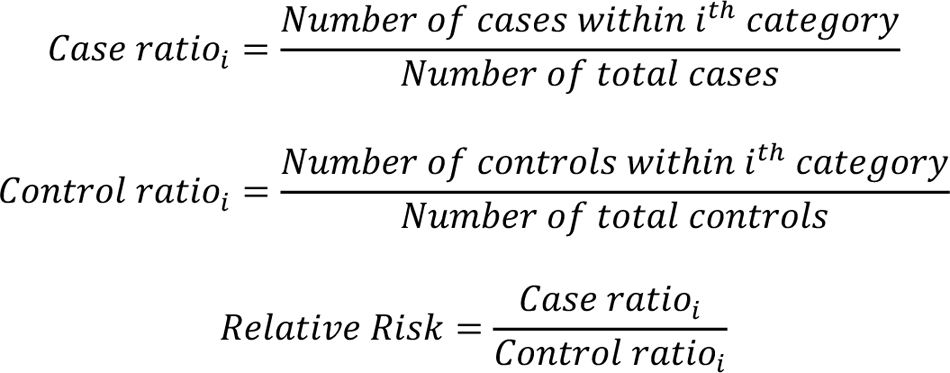

The binomial p-value is calculated by comparing two proportions: cases from total samples within a category and cases from total samples.

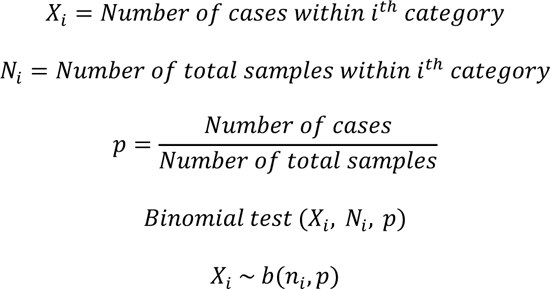

Case-control label swapping permutations generate p-values in both tests.

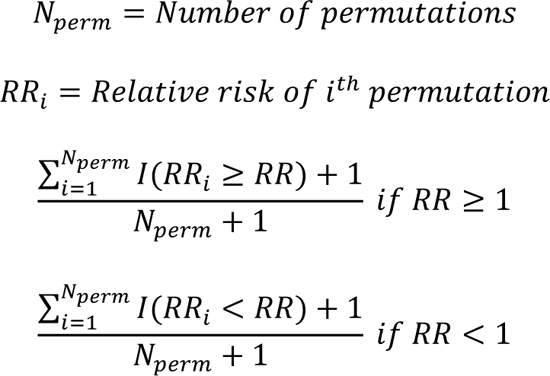

During burden tests, CWAS-Plus can correct confounding factors by adjusting the number of variants per sample. In this study, *de novo* variant counts were corrected for paternal age of birth.

### Risk score analysis

Risk scores for each category were generated to find the most effective predictors of the phenotype. We employed a Lasso regression model, focusing on rare categories (<2 variants in controls), and incorporating the number of variants within each category across samples as a metric. Approximately 80% of samples (n = 5,824 for variant-level analysis; n = 869 for sample-level analysis) constituted the training set, with the phenotype serving as the response variable. The optimal model was determined via the selection of the minimum lambda value from five-fold cross-validation, repeated 10 times. Model performance was evaluated using the R squared (R^2^) and significance was assessed through label-swapping 1,000 times, randomly assigning the phenotype to samples.

For feature selection, risk score analysis was applied to noncoding categories from each annotation dataset (i.e., gene set, functional annotation, functional score). Datasets with positive R^2^ values were further analyzed.

### Burden shift analysis

CWAS-Plus identifies overrepresented annotations from category-level burdens. In each permutation (n = 10,000), phenotypes are randomly assigned to samples while maintaining the original ratio. Subsequently, CWAS-Plus compares the number of significant categories of interest with those from each permutation, obtaining p-values for each phenotype.

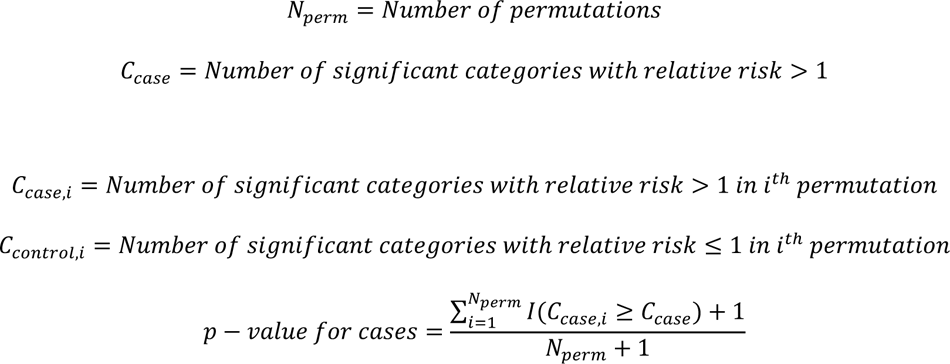

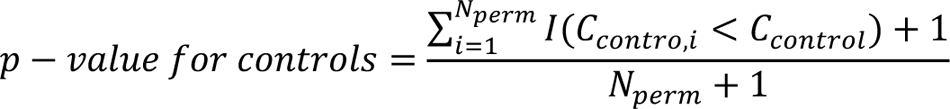

### Clustering of categories

CWAS-Plus clusters categories by calculating correlation values between two categories. The correlation is calculated based on the principle that under the null hypothesis the covariance is equivalent to the variance of shared variants^26^. Because a binomial random variable can be expressed as the sum of independent Bernoulli variables, the variance can be computed using the equation:

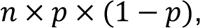

where n represents the number of tests and p denotes the binomial probability.

Each variant is treated as an independent test, replacing the total number of tests (n) with the number of variants. Consequently, the covariance between two categories is determined by the number of shared variants. We obtained a correlation matrix of size *c* × *c* by calculating the standardized covariance for all pairs of categories among the total.

### Finding the number of effective tests for burden test

To estimate the threshold for genome-wide significance, we converted the correlation matrix into a negative Laplacian metric, normalizing the value with the degree of each node. Eigen decomposition was then applied to identify the number of eigenvalues explaining at least 99% of the variance, which determined the effective number of tests.

### Detecting Association With Networks (DAWN) analysis

To investigate the subnetwork within noncoding risk categories, we employed the DAWN algorithm^27^. Correlation metrics or eigen vector metrics from risk-associated categories were transformed into two-dimensional coordinates via dimension reduction. Subsequently, coordinates were clustered using k-means or leiden clustering, and disease association was determined using a hidden Markov random field model. P-values and RRs for each category within clusters, as well as those of nearby clusters, were considered to compute z-scores and adjusted p-values. Variants from significant clusters were defined as risk noncoding variants.

### Examples for CWAS runs

For benchmarking CWAS-Plus, we used 255,106 *de novo* variants identified in 1,902 ASD families from our previous WGS study^24^.

For variant-level CWAS analyses, we obtained project VCF with joint genotyping in 4,270 ASD families from the Simons Simplex Collection (SSC)^28^ and Simons Foundation Powering Autism Research for Knowledge (SPARK)^29^. ASD families include 4,354 ASD cases and 2,926 unaffected control siblings. Data access and analyses were approved by the Institutional Review Board of Korea University (approval number: KUIRB-2022-0409-03). After quality control, 474,788 *de novo* variants identified from 7,280 samples were utilized (see ‘High quality variants for variant-level analyses’ section in **Supplementary Materials** for detailed methods).

For sample-level CWAS analyses, we obtained 63,667,178 variants from 1,196 individuals in the ROSMAP study^30,31^. After quality control, 25,052,701 variants from 1,087 samples were utilized (see ‘High quality variants for sample-level analyses’ section in **Supplementary Materials** for detailed methods).

### Downstream analysis

#### Transcription factor enrichment

We investigated enrichment of human transcription factors (n = 1,622)^32^ in genes affected by RNVs, PTVs, and noncoding variants (excluding RNVs). Fisher’s exact test, followed by false discovery rate (FDR) correction, determined significance with an adjusted p-value threshold of < 5.0×10^-2^. Background gene sets included all genes from GENCODEv44 and genes annotated to variants.

#### Correlation between DAWN clusters and single annotation datasets

To assess the correlation between DAWN clusters and single annotation datasets, we extracted variants from each cluster and annotation dataset. Correlation was calculated using the same method as in CWAS-Plus, where we determined the number of shared variants between two sets (either from clusters or single annotations), considering the number of variants in each set.

## RESULTS

### Overview of CWAS-Plus pipeline

The CWAS-Plus package includes several steps to explore noncoding associations of genetic disorders (**Figure 1A**). CWAS-Plus utilizes various datasets to annotate variants and extract relevant genomic information (**Figure 1B**). The annotated information is employed for variant categorization, in which categories are defined by the combination of five features: variant type, genomic region, gene set, functional annotation, and functional score (**Figure 1B**). Subsequently, association tests are conducted for each category (**Figure 1C**). CWAS-Plus utilizes these associations from single categories to provide more comprehensive measurements with multiple categories, enhancing noncoding association to identify variants contributing to disease risk more effectively.

**Figure 1.**
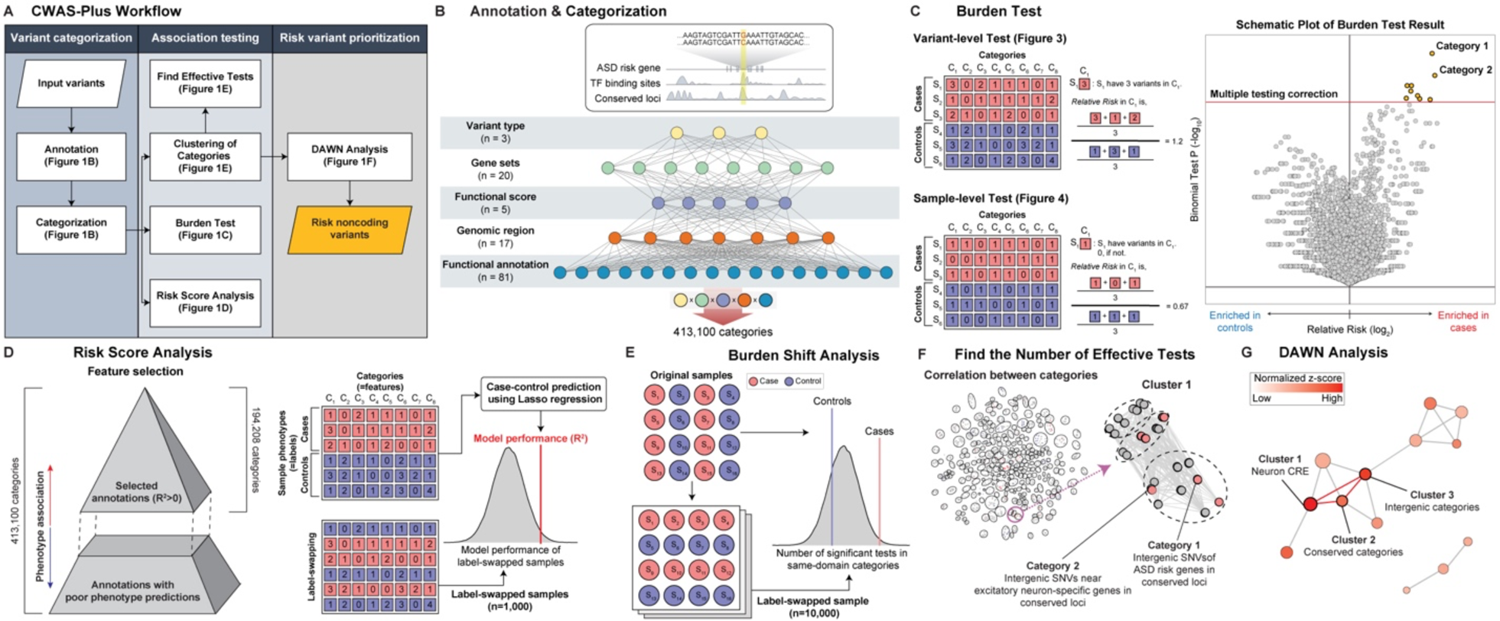
Overview of CWAS-Plus. **A.** Overview of the CWAS-Plus workflow. **B.** An example of annotation and categorization. Input variants are annotated and categorized based on five types of variant annotations. The combinations of domains result in a single category. **C.** Burden test for a variant- and sample-level test: the number in the boxes indicate the number of variants in each category across samples. The volcano plot contains the burden test result for single categories. The x-axis indicates the relative risk in the log_2_ scale, whereas the y-axis indicates two-sided binomial p-values in the log_10_ scale. Red dashed line indicates the study-wide significance threshold for the number of effective tests. **D.** Risk score analysis: through feature selection, annotation datasets with high phenotype association are selected for further analyses. Label-swapping randomly assigns phenotypes to samples. The gray density plot displays R^2^ distribution of label-swapped samples (n = 1,000). The red line represents observed R^2^ from original samples. **E.** Burden shift analysis: Label-swapping randomly assigns phenotypes to samples. The circles and colors represent samples and phenotypes, respectively. Density plot illustrates the distribution of significant category counts for label-swapped samples. The observed number of significant categories is depicted as lines in the plot. **F.** Find the number of effective tests: Categories are clustered based on pairwise correlations to find the number of effective tests. Each dot represents a category, and black circles denote clusters. The purple arrow highlights a representative network subset, with color indicating burden direction in each category. **G.** DAWN analysis: The network illustrates the relationship of clusters, with node color indicating the degree of disease association (z-score scale) and node size reflecting cluster size (number of categories).

With categorized variants, risk score analysis identifies categories that predict disease phenotype (**Figure 1D**). The predictive contribution of each category is assessed by a Lasso regression model using the category patterns of each sample. In risk score analysis, feature selection is conducted to select phenotype-relevant datasets with positive model performance (R^2^) for further analyses. Subsequently, burden shift analysis identifies overrepresented annotations in category-level association tests (**Figure 1E**). The significant excess of association compared to the null distribution suggests disease risk. Comparing results from both analyses assesses signal concordance, yielding candidate categories for disease association and subsequent Detecting Association With Networks (DAWN) analysis. Furthermore, CWAS-Plus provides a reliable method (**Figure 1F**) to determine the number of effective tests. By treating highly correlated categories as a single effective test, it facilitates accurate estimation of the study-wide significance. In DAWN analysis, categories are clustered and investigated for associations with disease risk (**Figure 1G**). Based on the DAWN algorithm^27^, CWAS-Plus constructs a network of clusters and evaluates disease risk considering nearby clusters, enhancing the identification of variants with risk.

### Significant clusters yield risk noncoding variants (RNVs), potential pathogenic variants with noncoding associations

The CWAS-Plus pipeline, described above, receives a list of variants as input and comprises seven steps to assess the noncoding association. We performed variant-level analysis using *de novo* variants from 7,280 samples in ASD families. Additionally, sample-level analyses utilized rare variants from 1,087 WGS samples in the Religious Orders Study and the Memory and Aging Project (ROSMAP)^30^.

### Performance improvements in CWAS-Plus

CWAS-Plus offers several advantages over CWAS^24^ (**Table 1**), owing to its transformation into a user-friendly Python package with simplified source code. Researchers can easily employ its capabilities through straightforward parameter settings. The reorganization of the source code improves comprehensibility as well as enables streamlined multiprocessing, resulting in shorter processing times.

**Table 1.** Comparison of CWAS-Plus and CWAS.

|  | CWAS | CWAS-Plus |
| --- | --- | --- |
| RAM | ≥16 GB | ≥16 GB |
| Total execution time | 142 hours | 170 minutes |
| Programming language | R, Python2 | Python3 |
| Packaging | No | Yes |
| Multiprocessing | Yes | Yes |
| Batching | No | Yes |
| Setting environment | n/a | Pip |
| Annotation harmonization support | No | Yes |

Both tests were conducted using 36 cores and 256 GB of memory.
n/a, not applicable

We benchmarked the performance of CWAS-Plus and compared a run time between CWAS-Plus and CWAS^24^. We performed the CWAS analysis for *de novo* variants of ASD families using the CWAS-Plus package and our previous CWAS scripts (https://github.com/sanderslab/cwas). For this, we obtained the annotation (total 27,462 categories) and sample dataset (1,902 ASD cases and 1,902 unaffected controls) from the previous study^24^ and evaluated the comparison in the same computing resource (36 CPU threads and 256 GB of memory). CWAS-Plus was 50 times faster than that of CWAS, highlighting its superior computational speed (Figure 2A, **Supplementary Table 1**).

**Figure 2.**
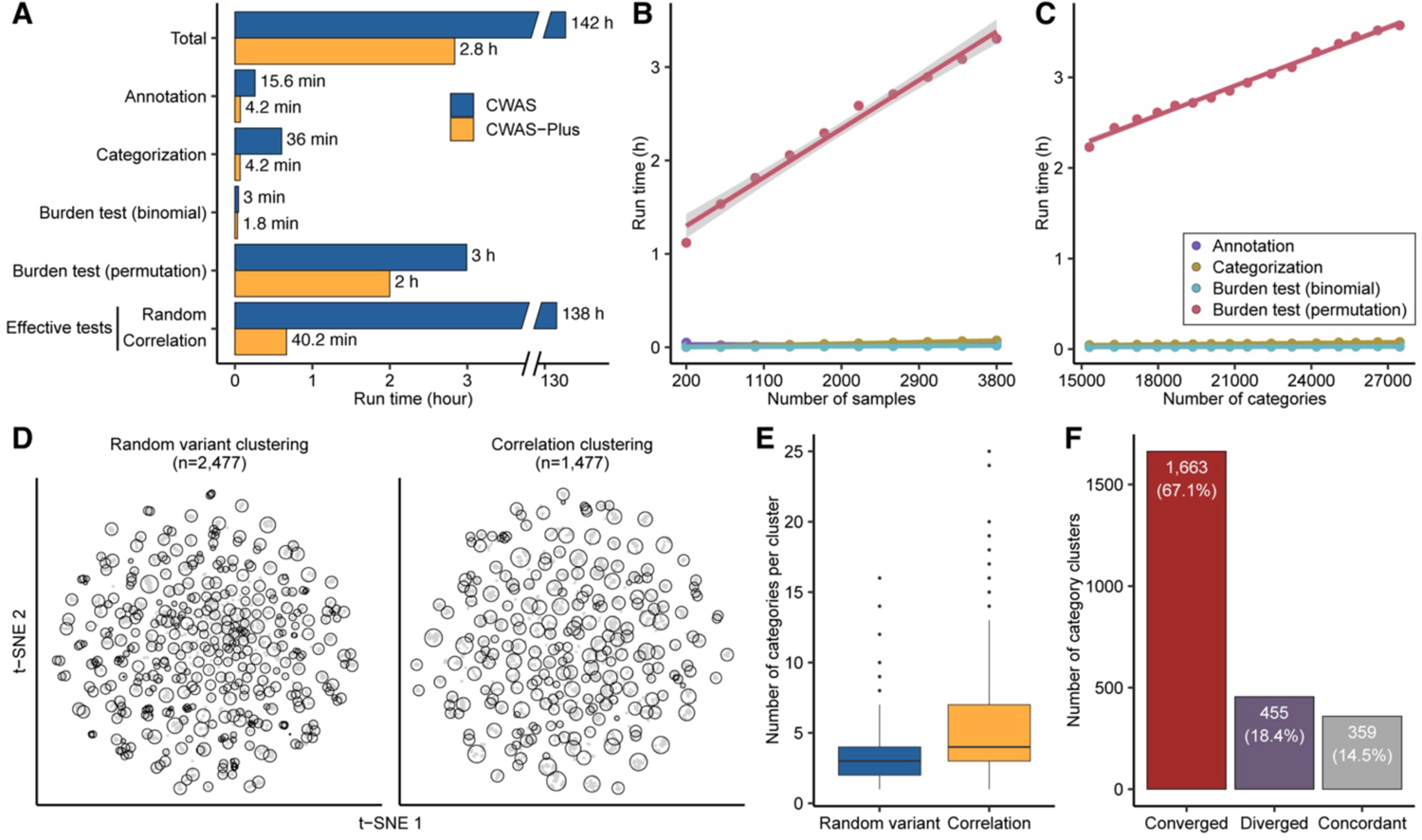
Benchmarking of CWAS-Plus. **A.** Comparison of run time between CWAS^24^ and CWAS-Plus. Run time was measured using the same annotation datasets, along with equivalent computing resources. The y-axis indicates each step of CWAS-Plus. The x-axis represents the run time (hours). The numbers next to bars refer to run time of each step (h for hour; min for minute). The comparison was conducted on 3,804 samples and 27,462 categories. **B.** Regression line illustrating the relationship between execution time and sample size increment. The dots indicate the observed time point, and the color indicates each process in CWAS-Plus. The gray shade represents the confidence interval. **C.** Regression line depicting the relationship between execution time and the number of category increment. The dots indicate the observed time point, and the color indicates each process in CWAS-Plus. The gray shade represents the confidence interval. **D.** t-SNE plots displaying the clustered features obtained from random variant clustering and correlation clustering methods. **E.** Distribution of the number of categories within each cluster using random variant clustering and correlation clustering methods. The y-axis indicates the number of categories per cluster. **F.** Differentiation of clusters from random variant clustering (random variant cluster) to clusters from correlation clustering (correlation cluster). The y-axis represents the number of random variant clusters, while the x-axis indicates the types of differentiation: convergence, divergence, and concordant. Convergence and divergence correspond to whether the random variant clusters were merged into a larger cluster or split into smaller clusters in correlation clusters, respectively. Concordant denotes instances where a random variant cluster aligns with a correlation cluster. **A-C.** Overall application of CWAS and CWAS-Plus was performed with 36 CPUs and 256GB RAM.

We further assessed the computational efficiency of CWAS-Plus by examining its CPU and memory usage. We observed that CWAS-Plus operates efficiently with a single CPU, leading to reasonable execution times. For datasets with <27,000 categories, CWAS-Plus was sufficient with <16 GB of RAM, highlighting efficient memory usage. The scalability of the CWAS-Plus package was tested for run with an increasing number of samples (Figure 2B, **Supplementary Table 2**) and datasets (Figure 2C, **Supplementary Table 3**). The results revealed a linear increase in execution time with each additional 400 samples, requiring 15 minutes. Similarly, adding a new functional annotation increased the time by approximately 6 minutes. These findings show that the speed of the CWAS-Plus depends on the number of samples and categories. Notably, CWAS-Plus can handle over 27,000 comparisons with approximately 4,000 samples in 3 hours, highlighting its efficiency.

CWAS-Plus also provides an efficient approach to find the number of effective tests for multiple testing correction. To accomplish this, CWAS^24^ utilized 10,000 sets of random variants. The p-values of the association tests, calculated from each variant set, were then used to measure the correlation between tests (categories). Considering that each variant set requires the same amount of time and memory resources for a single CWAS-Plus execution, this analysis incurs substantial computational costs.

Regarding this issue, CWAS-Plus offers a correlation-based method, saving substantial time compared to the former. Both methods cluster categories based on correlation values, yet the correlation-based method calculates the correlation using the number of variants (or samples, in sample-level analysis) shared between two categories. Therefore, the method is fast and effective.

For validation, we compared the outcomes from both methods (Figure 2D). As a result, we observed that the number of clusters in random variant clustering (n = 2,477) was greater than that in correlation clustering (n = 1,477). Additionally, we found a higher prevalence of larger clusters (more categories per cluster) in correlation clustering than in random variant clustering (Figure 2E). These observations collectively suggest that small clusters in random variant clustering may merge into a single cluster within the correlation clustering approach.

We then examined whether multiple clusters from random variant clustering (random variant cluster) converged to a larger cluster in correlation clustering (correlation cluster). Quantifying cluster similarities identified three scenarios: converged, diverged, and concordant. The converged clusters, wherein multiple random variant clusters merged into a single correlation cluster, constituted 67.1% of all random variant clusters (Figure 2F). Conversely, the diverged clusters, in which a random variant cluster split into multiple correlation clusters, accounted for 18.4%. This observation implies that, in most instances, random variant clusters converged toward a single correlation cluster.

Taken together, the results support that the number of correlation clusters is smaller than random clusters because random clusters merge into correlation clusters. However, the composition of categories within each cluster remains fairly similar, as correlation clusters absorbed small random clusters. Overall, our results demonstrate that the correlation-based method effectively replaces the utilization of random variant sets, considerably reducing computational time and memory usage.

### Application of CWAS-Plus to regulatory association in cell-type-specific functional data

Ongoing efforts have been made to generate various annotation datasets for functional regions in the noncoding genome or regulatory enhancers of various cell types and tissues. CWAS-Plus facilitates CWAS analysis by incorporating a new annotation dataset and seeking an appropriate multiple comparison level. Here, we performed the CWAS analyses for *de novo* variants in ASD cases (n = 4,354) and their unaffected siblings (n = 2,926) and examined noncoding associations of ASD in cell-type-specific regulatory elements. For this, we obtained various functional datasets, including single-nucleus ATAC-seq (snATAC-seq) datasets^3,33–35^ from early developmental stages of the human brain (a total of 111 annotation datasets) (Figure 3A).

**Figure 3.**
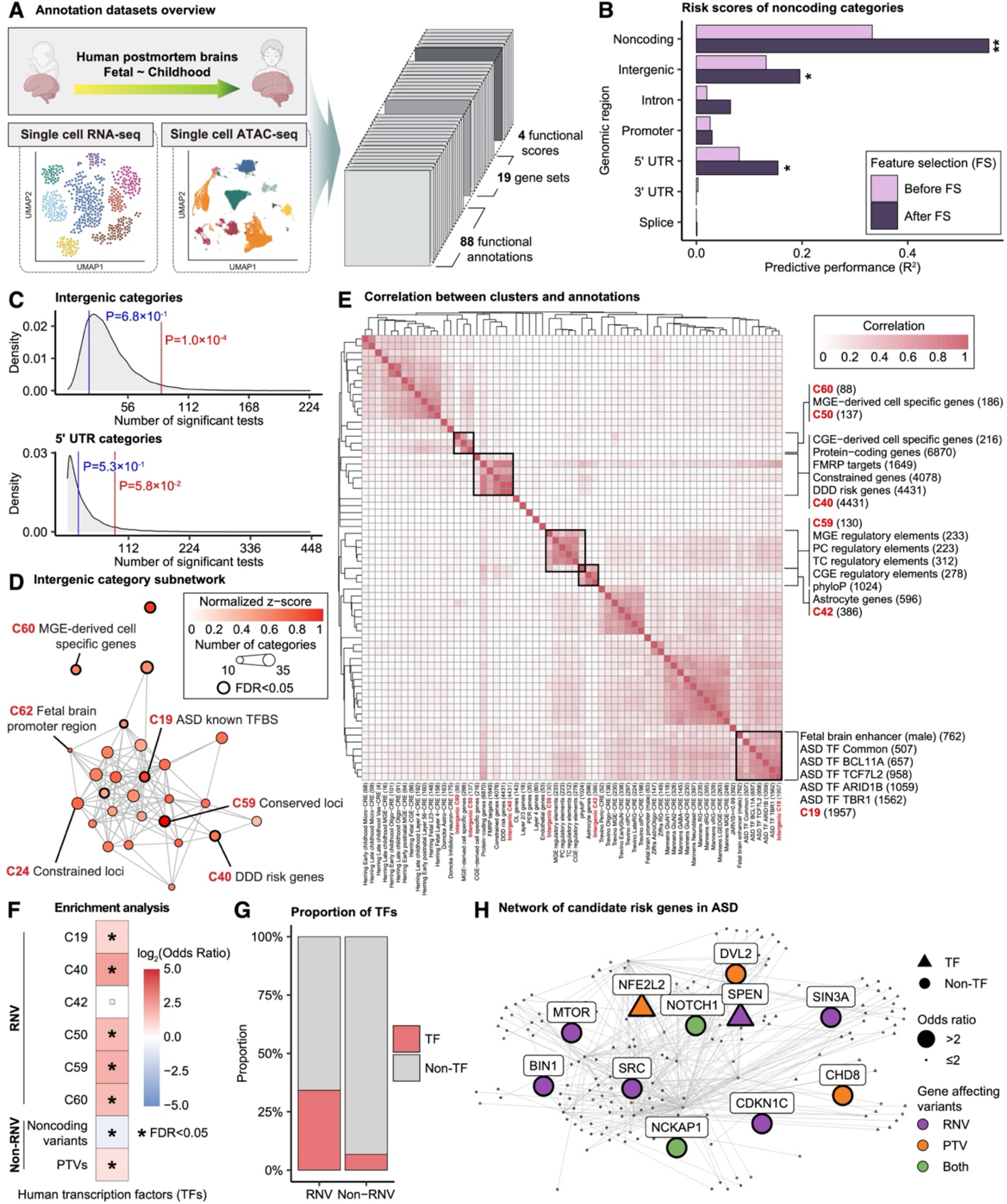
Noncoding association with ASD in cell-type-specific regulatory elements **A.** Summary of annotations for the CWAS analysis for ASD. Created with BioRender.com. **B.** Risk score analysis results of noncoding genomic regions. The x-axis indicates the model performance (R^2^) obtained from the Lasso regression model. The asterisks indicate permutated p-values (*, P < 5.0 ×10^-2^; **, P < 1.0×10^-2^). The color of the bars show whether feature selection is applied. **C.** Burden shift analysis results for intergenic and 5’ UTR categories. The gray density plots indicate the null distribution, and the vertical lines refer to observed number of significant tests for cases (red) and controls (blue). **D.** DAWN analysis results with intergenic categories. Each dot represents a single cluster. The degree of disease association is shown by color of clusters scaled with normalized z-scores. The node size indicates the number of categories within a cluster. The edge refers to the p-value correlation between clusters. The thick borderline indicates clusters with FDR < 5.0×10^-2^. **E.** Correlations between clusters and single annotations. The degree of correlations is shown by color. Thick boxes highlight annotations highly correlated with clusters. **F.** Enrichment analysis results with transcription factors. The asterisks indicate significance (FDR < 5.0×10^-2^). The color of boxes shows the degree of odds ratio. **G.** Comparison of the percentage of transcription factors in genes affected by RNVs and non-RNVs. The color represents the portion of transcription factors (red) and others (gray). **H.** Protein-protein-interaction network of genes in transcription regulation pathway and also carry RNV or PTV. The shape of nodes indicates whether the gene is transcription factor (triangle) or not (circle). The size of nodes shows the degree of odds ratio. The color of nodes represents the type of variants the gene carries.

Our CWAS analysis generated 142,498 categories for burden testing. As none of the noncoding categories exhibited study-wide significance (P < 1.4×10^-5^), we applied risk score analysis to measure noncoding association from multiple categories of each genomic region. The results showed that noncoding, including intergenic (R^2^ = 0.20%, P = 1.3×10^-2^; **Supplementary Table 4, 5**) and 5’ UTR (R^2^ = 0.15%, P = 1.1×10^-2^), categories were significantly enriched (Figure 3B). The performances were prominent after feature selection, indicating its necessity for enhancing model performance. We also performed burden shift analysis and found case-enrichment in intergenic categories (P = 1.0×10^-4^), supporting the association found from risk score analysis (Figure 3C).

We further explored intergenic variants through DAWN analysis, aiming to identify subnetworks within categories and find risk variants. Leveraging the correlations between categories, a network of intergenic categories was constructed (Figure 3D). We identified eight clusters associated with ASD (FDR < 5.0×10^-2^). Clusters of constrained loci, conserved loci, and ASD-associated transcriptional regulator binding sites were densely connected, offering insights into potential regulatory connections governing these loci.

To examine cluster characteristics, we identified annotation datasets correlated with the clusters. The results revealed cell-type-specificity in clusters 50, 60 and 42, where the former two clusters were highly correlated with medial ganglionic eminence-derived cells and the latter correlated with astrocytes (Figure 3E). In contrast, cluster 19 was specific to binding sites targeted by ASD-associated transcriptional regulators^36^. The results also provided shared features among datasets at the variant level. Specifically, excitatory neuron-cis-regulatory elements (CREs) in early stages were grouped, suggesting more specificity compared to late stages.

We defined variants within significant clusters as RNVs. To validate our results, we utilized deep-learning prediction tool, Sei^37^, to assess the regulatory activities of RNVs. Our observations revealed that variants from most clusters exhibited a higher percentage of variants in higher score bins than non-RNVs (**Supplementary Figure 1**), indicating that CWAS-Plus prioritized more pathogenic variants within the broader pool of noncoding variants.

While investigating the role of RNVs in ASD, we hypothesized that RNVs may not only affect the binding sites of transcription factors, but also affect transcription factors themselves, thereby disrupting transcriptional regulation. We utilized a list of human transcription factors^32^ to conduct enrichment analysis on both RNV-affected and non-RNV-affected genes. The results demonstrated significant enrichment in RNV-affected genes across most clusters compared to that in other noncoding variants (Figure 3F). Additionally, the transcription factors^32^ showed five-fold enrichment in RNVs compared to non-RNVs (Figure 3G), indicating the disruptive influence on gene expression through interference with transcription factor activity.

To understand the regulatory network affected by RNVs, we constructed a protein interaction network involving genes affected by both RNVs and protein-truncating variants (PTVs) (Figure 3H). This network specifically focused on genes involved in the transcription regulation. Genes with a high odds ratio (OR) for carrying *de novo* variants in ASD cases (OR > 2) showed associations with neurodevelopmental risk. For example, *NCKAP1*, crucial for neuronal differentiation, is implicated in ASD and neurodevelopmental delay^38,39^. Additionally, *CDKN1C*, exclusively pinpointed by RNVs, is previously reported as a neurodevelopmental risk gene^39^. These findings support the reliability of our analyses, providing insights into potential associations of risk genes regulated by noncoding variants.

### Application of CWAS-Plus to sample-level analysis of rare variants associated with Alzheimer’s disease

While the CWAS framework was mainly developed to test noncoding association for *de novo* variants, it can be applied to rare variants^23^. For burden tests, the CWAS-Plus package provides a “sample-level test”, estimating case-control association by comparing the number of samples carrying rare variants in each category (Figure 1C). Unlike *de novo* variants, the number of rare variants is highly variable across individuals due to different genetic backgrounds, potentially leading to spurious genetic associations with the variant-level test. Thus, the CWAS analysis for rare variants should be performed by the sample-level test to yield robust signals.

We evaluated the performance and reproducibility of the sample-level test results against the variant-level test using *de novo* variants. CWAS analyses for *de novo* variants showed highly consistent estimates between the variant-level and sample-level tests (**Supplementary Figure 2, Supplementary Table 6**). Given these results, we infer that sample-level tests similarly capture associations to variant-level tests.

With the objective to demonstrate the ability of CWAS-Plus to assess noncoding risk in other genomic disorders, we applied the sample-level test to rare variants in 734 Alzheimer’s disease (AD) cases and 353 controls from ROSMAP WGS data. Annotation datasets included AD-specific CREs^40^ and differentially expressed genes from single-cell data^41^. Among 55,445 categories, none reached study-wide significance (P < 4.9×10^-5^). Since no single category fully explained AD risk associated with rare noncoding variants, we conducted risk score analysis to identify predictors among diverse categories (Figure 4A**, Supplementary Table 7, 8**). Noncoding categories, including intergenic (R^2^ = 1.26%, P = 4.3×10^-2^), intron (R^2^ = 1.67%, P = 3.8×10^-2^), and 3’ UTR (R^2^ = 1.51%, P = 7.0×10^-3^), were significant, suggesting potential AD risk in these categories.

**Figure 4.**
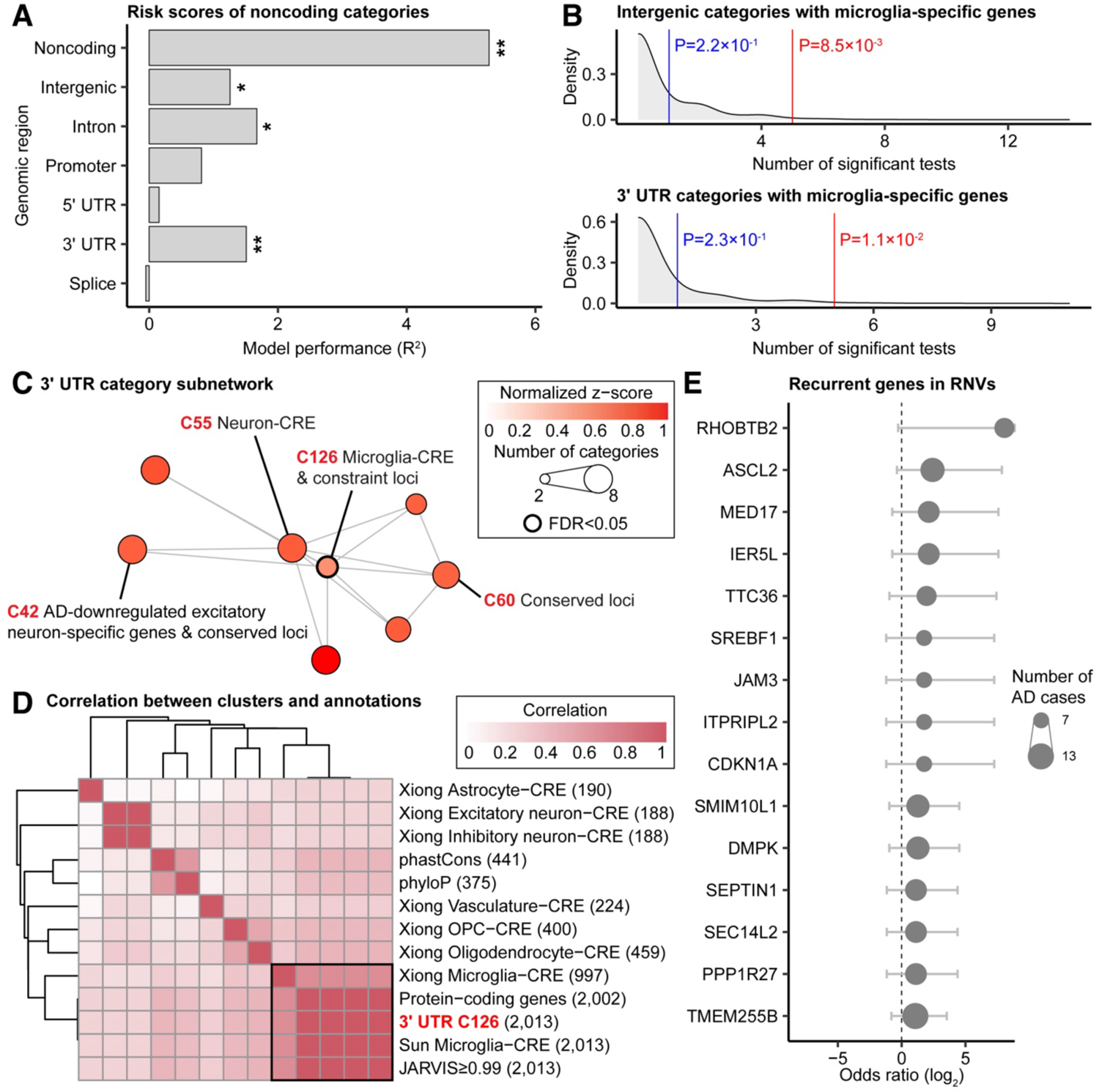
Application of CWAS-Plus to rare variants in Alzheimer’s disease **A.** Risk score analysis results of noncoding genomic regions. The x-axis indicates the model performance (R^2^) obtained from the Lasso regression model. The asterisks indicate permutated p-values (*, P < 5.0×10^-2^; **, P < 1.0×10^-2^). **B.** Burden shift analysis results for intergenic and 3′ UTR categories with microglia-specific genes. The gray density plots indicate the null distribution, and the vertical lines refer to observed number of significant tests for cases (red) and controls (blue). **C.** DAWN analysis results with 3′ UTR categories. Each dot represents a single cluster. The degree of disease association is shown by color of clusters scaled with normalized z-scores. The node size indicates the number of categories within a cluster. The edge refers to the p-value correlation between clusters. The thick borderline indicates clusters with FDR < 5.0×10^-2^. **D.** Correlations between clusters and single annotations. The degree of correlations is shown by color. **E.** Genes recurrent in RNVs. The x-axis indicates the odds ratio in log_2_ scale. The circle size shows the number of AD cases.

We then applied burden shift analysis, confirming case-enrichment in intergenic and 3′ UTR categories for microglia-specific genes (Figure 4B). The microglia signals underscores the robustness of CWAS analysis, given the well-established disruption of neuroinflammatory pathways in AD pathology^42^.

Focusing on 3′ UTR variants, we applied DAWN analysis and found one significant cluster (cluster 126; FDR < 5.0×10^-2^), defining variants in the cluster as RNVs for AD (Figure 4C). To delineate the distinctive features of this cluster, we identified single annotations highly correlated with the cluster (Figure 4D). The cluster showed enrichment with microglia-specific CREs and constrained loci, indicating the regulatory roles of RNVs in microglia-specific pathways.

We evaluated gene recurrence in 526 cases with RNVs, identifying 41 genes in more than five AD cases (OR > 1), providing a candidate list for AD risk genes regulated by RNVs (Figure 4E). For example, *CDKN1A* (OR = 3.39; 95% confidence interval (95% CI): 0.4–153.1), a cyclin-dependent kinase inhibitor and a senescence marker associated with the p53 signaling pathway^43–45^, is a potential regulator of cell cycle progression in AD pathogenesis.

Additionally, five genes (i.e., *CDK10*, *KLF16*, *SNX1*, *SORT1*, and *TSPAN14*) overlapping with AD risk genes from a recent GWAS results^46^ were identified. Regarding KLF16, seven individuals (5 cases and 2 controls; OR = 1.2, P = 1; 95% CI: 0.2–12.7) carried RNVs, with four cases sharing the same variants (g.chr19:1853596G>A, hg38). These findings suggest an association between rare and common variant-targeted genes in AD, supporting the potential of CWAS-Plus in capturing reliable signals from rare variants.

### Utilizing large annotation datasets improves model performance in risk score analysis

One of the key questions in risk score analysis is whether adding more annotation datasets enhances the model performance. We compared three sets of annotation datasets: set 1, set 2, and set 3 (Figure 5A). Set 3 corresponds to the initial annotation dataset from Figure 3A (286 functional annotations and scores; 142,498 categories). Set 1 and 2 share the same datasets as set 3 but differ in functional annotations. Set 1 includes CREs from Herring *et al*.^3^ (20 functional annotations and scores; 48,680 categories), while set 2 expands from set 1 by adding regulatory elements from the Roadmap Epigenomics project^47^ and VISTA^48^, and binding sites for ASD-associated transcription regulators^36^ (274 functional annotations and scores; 69,702 categories).

**Figure 5.**
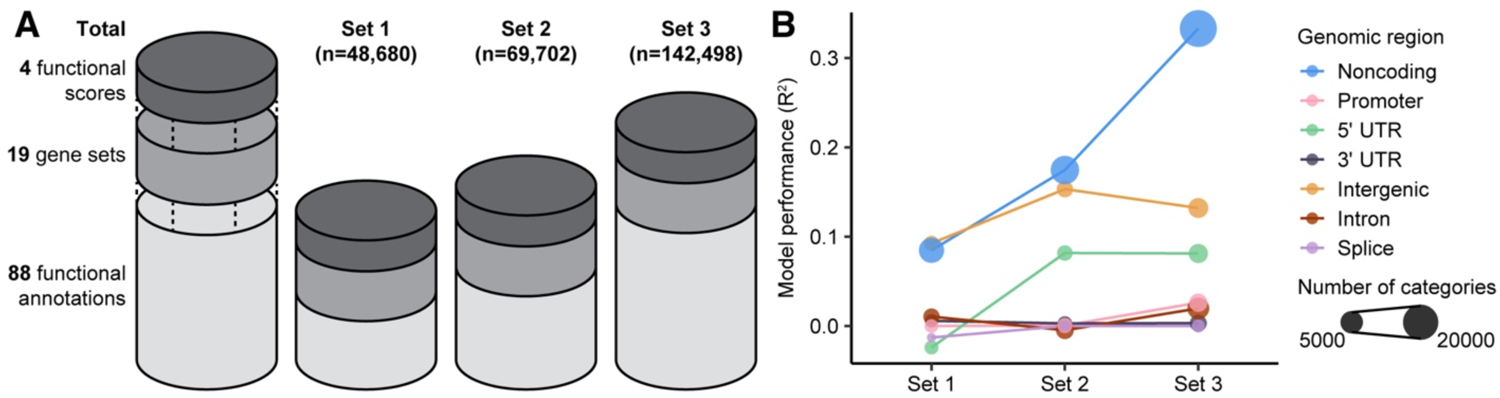
Improved model performance with expanded annotation datasets **A.** A schematic view of three annotation datasets with increasing size. The number of categories is denoted in each set. **B.** Model performance of categories from each genomic region in each set from **A**. The colors correspond to genomic regions, and the size of data points reflects the number of categories.

The introduction of additional regulatory elements improved the model performance within noncoding categories. Set 3 showed the highest performance, outperforming set 1, followed by set 2 (R^2^ = 0.08%, P = 5.5×10^-1^ for set 1; R^2^ = 0.17%, P = 4.3×10^-1^ for set 2; R^2^ = 0.33%, P = 3.7×10^-1^ for set 3; Figure 5B, **Supplementary Table 9**). While certain genomic regions, such as 5′ UTRs, exhibited improved model performance compared to set 1, others, like 3’ UTRs, demonstrated a decrease. These observations emphasize the importance of not only increasing the quantity of annotations but also the composition of the added features to enhance the performance.

## DISCUSSION AND CONCLUSION

In this study, we introduce CWAS-Plus, a tool for efficient CWAS analysis to identify noncoding associations from WGS data. The package supports user-friendly functions for categorization and burden enrichment test for various functional datasets. Our tool facilitates the identification of the number of effective tests across multiple tested categories for multiple testing correction. Consequently, it is applicable for WGS studies of various human disorders.

Here we integrated multiple functional datasets, including snATAC-seq datasets from human cortex, for CWAS analysis of *de novo* variants in ASD families, demonstrating cell-type-specific context for noncoding association. Significant associations were observed in intergenic categories enriched with regulatory regions of fetal brains and binding sites of ASD-associated transcription regulators. These results suggested that RNVs may contribute to ASD neurobiology by disrupting transcription factors during fetal brain development. Moreover, consistent with our previous finding^24^, RNVs were highly correlated with sequence conservation, indicating ASD noncoding association for regulatory elements with evolutionary conservation. Overall, CWAS-Plus enables integration of accumulated functional datasets with WGS data, thereby facilitating the identification of novel associations between regulatory elements and noncoding variants.

CWAS-Plus provides a novel approach to assess the genome-wide significance for rare noncoding variants. Unlike GWAS, which estimates effective association tests based on locus correlation, CWAS-Plus addresses the absence of standardized criteria for closely related association tests in rare variants by introducing a method to calculate associations between tests (categories). This unique methodology enables CWAS-Plus to conduct correction for multiple hypothesis testing, providing a tailored significance threshold for rare variants.

CWAS-Plus offers a comprehensive approach performing analyses at both the variant-level and sample-level. *De novo* variants occur at a relatively consistent rate across individuals. In contrast, rare variants show higher variability in occurrence rates due to genetic ancestry. CWAS-Plus considers these characteristics and provides both variant- and sample-level tests, allowing its application to *de novo* and rare variants effectively. Applying CWAS-Plus to rare variants of AD cohort, despite the limited sample size, revealed potential risks associated with 3′ UTR variants in microglia-specific CREs. These results are consistent with previous findings where microglia-expressed genes were enriched with candidate causal genes found in GWAS studies^46,49,50^. Taken together, our findings emphasize the reliability of CWAS-Plus and support the applicability to rare variants and complex genomic disorders, such as AD.

Despite various advantages, our package has a few limitations. CWAS-Plus enables users to customize annotation datasets for their phenotype of interest. However, the degree of freedom may pose challenges for diseases for which there is no access to relevant datasets or clear hypotheses for constructing customized categories. To address this issue, we provide datasets, such as putative promoter and enhancer regions, as a baseline to facilitate a more accessible starting point. Also, CWAS-Plus offers feature selection to select phenotype-relevant datasets, refining annotation dataset composition. Furthermore, CWAS-Plus has significantly improved computational speed; however, computation testing at a cohort level with hundreds of thousands of samples has not been carried out yet. Continuous development efforts will be undertaken to overcome these limitations of CWAS-Plus.

In summary, we present CWAS-Plus, a Python package for performing category-wide association tests for the genome-wide assessment of noncoding associations. CWAS-Plus offers an efficient and user-friendly approach for integrating functional datasets with large-scale WGS data and empowers multiple testing comparisons. Our package is applicable to both *de novo* and rare variants. With CWAS-Plus, we expect to uncover novel noncoding associations with rare variants and enhance our understanding of genetic contributions to pathologies.

## ETHICS APPROVAL AND CONSENT TO PARTICIPATE

Informed consent was obtained from all participants involved in the study by the Simons Foundation Autism Research Initiative (SFARI) and Religious Orders Study (ROS) and the Rush Memory and Aging Project (MAP). Data access and analyses were approved by the Institutional Review Board (IRB) of Korea University (approval number: KUIRB-2022-0409-03). The ROSMAP were approved by the IRB of Rush University Medical Center.

## AUTHOR’S CONTRIBUTIONS

Study design, Y.K., M.J., R.Y., D.W., K.R., B.D., S.S., and J.-Y.A.; Data processing, Y.K., I.G.K., and H.L; Data analysis, Y.K.; Package development, Y.K., M.J., I.G.K., J.H.K., S.S., and J.-Y.A.; Manuscript preparation, Y.K., I.G.K., and J.-Y.A.; Supervision, S.S., D.W., J.P., I.B.K. and J.-Y.A. All authors have read and approved the final draft of the manuscript for submission.

## Data Availability

The source code of CWAS-Plus is available via a GitHub repository (https://github.com/joonan-lab/cwas/) and Zenodo (https://doi.org/10.5281/zenodo.10795678)^51^, with a user manual (https://cwas-plus.readthedocs.io/en/latest/). Both repositories are released under the MIT license. The scripts to produce the annotation dataset, along with the burden test results from the CWAS analyses conducted in this study, are available via Zenodo (https://doi.org/10.5281/zenodo.10814080)^52^. The Zenodo repository is released under the Creative Commons Attribution 4.0 International license. CWAS-Plus is written in python (version≥3.9), and easy to install and use in Linux or Mac OS. *De novo* variants were obtained from the pVCF file, accessible with approval from the Simons Foundation Autism Research Initiative (SFARI Base; https://sfari.org/resources/sfari-base). ROSMAP WGS data can be requested at the AD Knowledge Portal under accession code syn10901595 (https://www.synapse.org/#!Synapse:syn10901595; see https://adknowledgeportal.synapse.org/Data%20Access for data access instructions).

## ACKNOWLEDGEMENTS

We are grateful to all of the families at the participating Simons Simplex Collection (SSC) sites, as well as the principal investigators (A. Beaudet, R. Bernier, J. Constantino, E. Cook, E. Fombonne, D. Geschwind, R. Goin-Kochel, E. Hanson, D. Grice, A. Klin, D. Ledbetter, C. Lord, C. Martin, D. Martin, R. Maxim, J. Miles, O. Ousley, K. Pelphrey, B. Peterson, J. Piggot, C. Saulnier, M. State, W. Stone, J. Sutcliffe, C. Walsh, Z. Warren, E. Wijsman). We appreciate obtaining access to phenotypic and genetic data on SFARI Base. Approved researchers can obtain the SSC population dataset described in this study by applying at https://base.sfari.org. We also thank to the participants in Religious Order Study, the Memory and Aging Project (ROSMAP). The results published here are in whole or in part based on data obtained from the AD Knowledge Portal (https://adknowledgeportal.org). Study data were provided by the Rush Alzheimer’s Disease Center, Rush University Medical Center, Chicago. Data collection was supported through funding by NIA grants P30AG10161 (ROS), R01AG15819 (ROSMAP; genomics and RNAseq), R01AG17917 (MAP), R01AG30146, U01AG32984 (genomic and whole exome sequencing), U01AG46152 (ROSMAP AMP-AD, targeted proteomics), U01AG61356 (whole genome sequencing, targeted proteomics, ROSMAP AMP-AD), the Illinois Department of Public Health (ROSMAP), and the Translational Genomics Research Institute (genomic). Additional phenotypic data can be requested at www.radc.rush.edu.

## AUTHOR BIOGRAPHIES

**Yujin Kim** is a graduate student at Korea University, where she focuses on studying noncoding variants that disrupt enhancers implicated in autism and developing a statistical method for the genetic association of rare variants from whole-genome sequencing data.

**Minwoo Jeong** worked as a research assistant with a particular interest in developing bioinformatic methods for large-scale whole-genome sequencing projects.

**In Gyeong Koh**, a graduate student at Korea University, is currently engaged in developing a single-cell sequencing atlas and data integration methods for various human brain transcriptome studies.

**Chanhee Kim** is a graduate student at Korea University, who is studying rare noncoding variants in Alzheimer’s disease using category-wide association techniques and applying them to large-scale whole-genome sequencing studies of Alzheimer’s disease.

**Hyeji Lee** is a graduate student at Korea University and focuses on studying rare inherited variants in autism and neurodegenerative disorders.

**Jae Hyun Kim**, formerly a graduate student at Korea University, conducted research on noncoding structural variants in autism using whole-genome sequencing datasets.

**Ronald Yurko** is an assistant teaching professor in the Department of Statistics & Data Science at Carnegie Mellon University. Dr. Yurko is interested in developing methods at the interface of inference and machine learning, particularly oriented towards problems in statistical genetics and genetic association studies.

**Il Bin Kim**, a professor at the Department of Psychiatry, CHA University School of Medicine, is dedicated to psychiatric genetics and the identification of promoter-enhancer interactions in the noncoding genome from a whole-genome sequencing study of Korean autism families (Kim et al., 2022, Molecular Psychiatry).

**Jeongbin Park**, an assistant professor at the School of Biomedical Convergence Engineering, Pusan National University, focuses on developing machine learning and probabilistic tools for various genomic studies, and has contributed to the development of various packages for single-cell analysis.

**Donna M. Werling**, an assistant professor at the University of Wisconsin-Madison, conducts research on genetic risk factors for autism and other neuropsychiatric conditions, as well as sex differences in neurobiology and disease risk. Dr. Werling has developed bioinformatics tools, including the category-wide association test, for large-scale whole-genome sequencing studies (Werling et al., 2018, Nature Genetics) and identified regulatory noncoding variants associated with human cortical development (Werling et al., 2020, Cell Reports).

**Stephan J. Sanders**, a professor at the Department of Paediatrics, University of Oxford, and the Department of Psychiatry and Behavioral Sciences, University of California, San Francisco, is renowned for his landmark studies on *de novo* variants in autism (Sanders et al., 2012, Nature), autism-associated genes (Sanders et al., 2015, Neuron), and noncoding risk mutations in autism (An et al., 2018, Science). **Joon-Yong An**, an associate professor in the Department of Biosystem and Biomedical Science at Korea University, has been dedicated to autism genetics and large-scale whole-genome sequencing analysis. Dr. An introduced the category-wide association test method for whole-genome sequencing studies (Werling et al., 2018) and identified noncoding risk mutations in autism (An et al., 2018, Science).

## SUPPLEMENTARY FIGURES

**Supplementary Figure 1.**
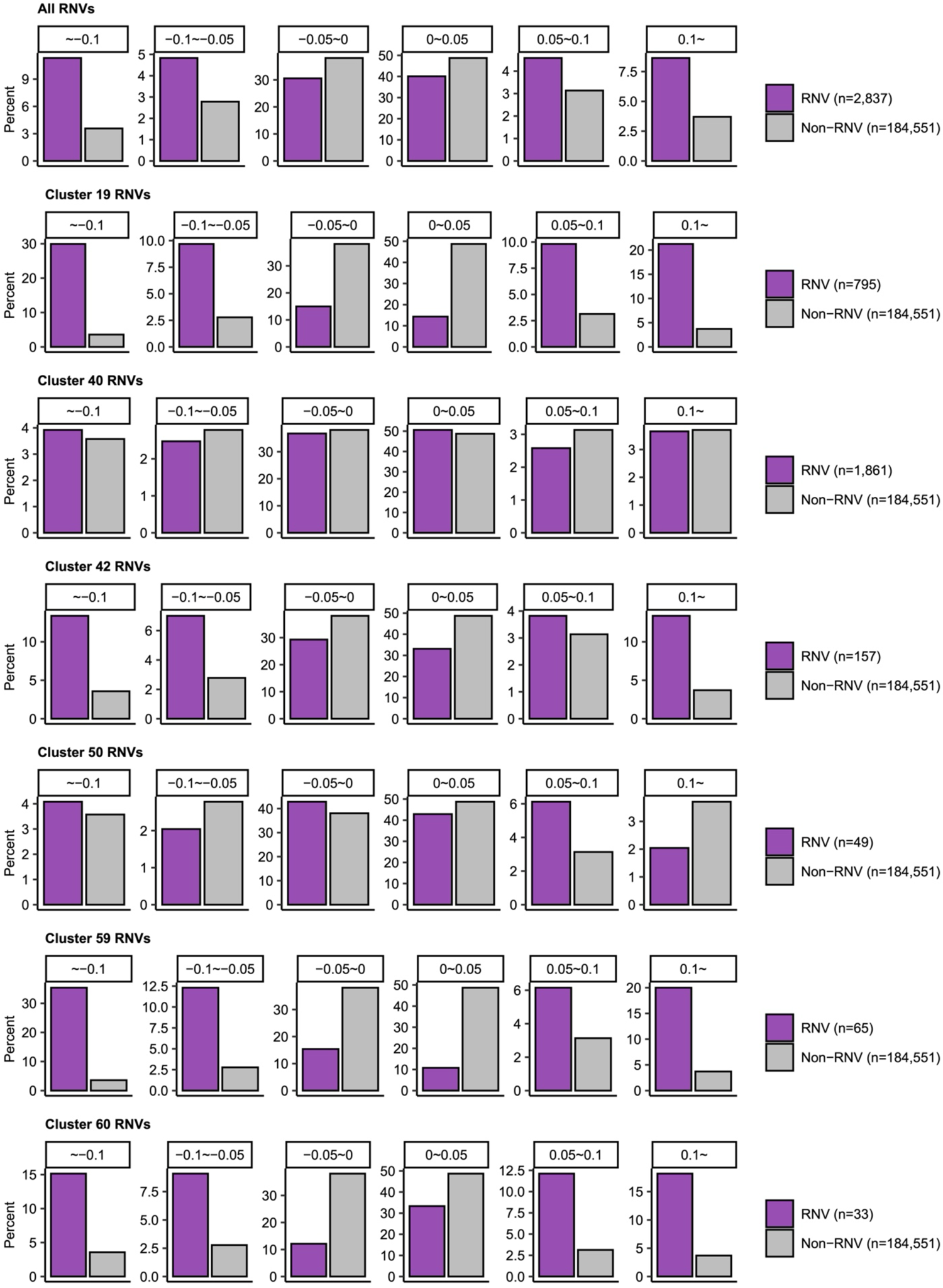
Deep-learning predictions of RNVs and non-RNVs using Sei Comparisons of the percentage of variants in specific range of deep-learning predictions estimated using Sei. The purple color refers to RNVs and gray color refers to the remaining intergenic variants.

**Supplementary Figure 2.**
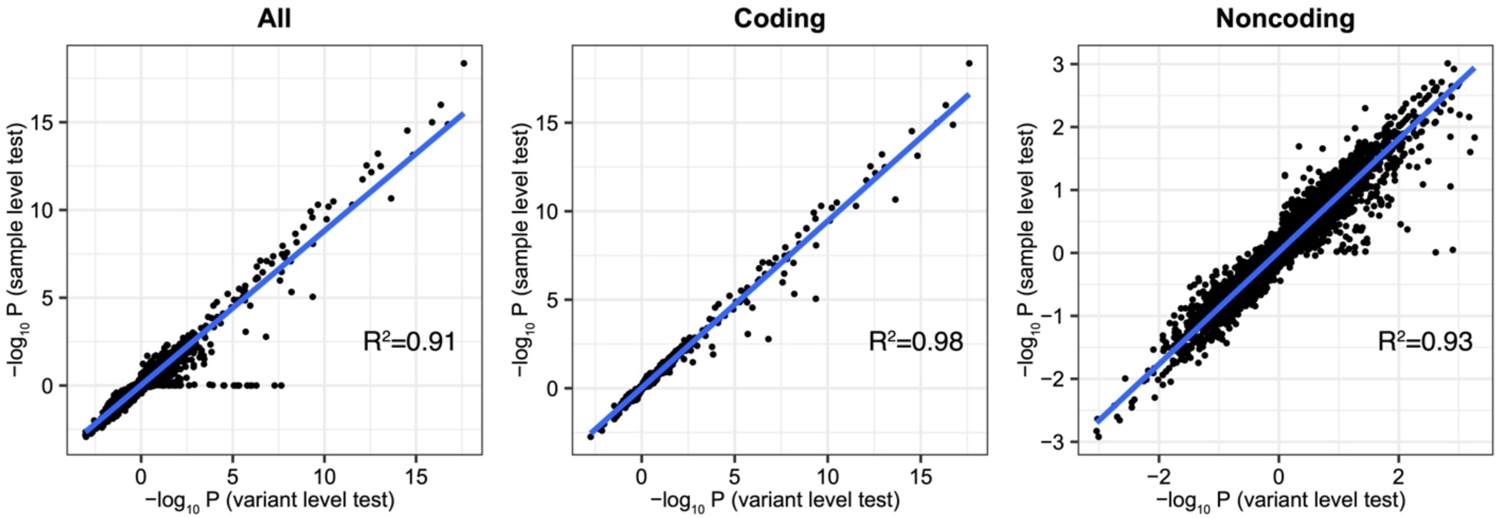
Comparison between variant-level and sample-level CWAS analyses. Correlations of burden test estimates between variant-level and sample-level tests with *de novo* variants. Correlation coefficients were measured using two-sided binomial p-values of categories from each test.

